# A Projection Model of COVID-19 Pandemic for Belgium

**DOI:** 10.1101/2020.05.31.20118406

**Authors:** Michael Ruzhansky, Niyaz Tokmagambetov, Berikbol T. Torebek

**Affiliations:** Department of Mathematics: Analysis, Logic And Discrete Mathematics Ghent University, Belgium and School of Mathematical Sciences Queen Mary University of London United Kingdom *E-mail address*; Department of Mathematics: Analysis, Logic and Discrete Mathematics Ghent University, Belgium and Al–Farabi Kazakh National University Almaty, Kazakhstan *E-mail address*; Department of Mathematics: Analysis, Logic and Discrete Mathematics Ghent University, Belgium and Institute Of Mathematics And Mathematical Modeling, Almaty, Kazakhstan *E-mail address*

## Abstract

We consider a simple model for the COVID-19 pandemic to analyse the relative effectiveness of several stages of the lockdown in Belgium, as well as of several phases of its relaxation. We also make a future projection of different types of measures relative to different stages of the already experienced lockdown.

## 1. Introduction

Starting in December 2019 in Wuhan, the severe acute respiratory syndrome coronavirus 2 (SARS-CoV-2) has been spreading around the world, causing the coronavirus disease 2019 (COVID-19). For everyday statistics, we refer to the site [1], the data from which we used for our modelling. The first case in Belgium was detected on 4 February 2020, and the number of all registered infected people reached 58381 by 31 May 2020.

Belgium has already experienced a lockdown with subsequent relaxation phases of different strengths. The effective lockdown in Belgium was introduced on 13 March 2020, and the official one from 18 March 2020. Subsequently, Phase Ia took effect on 4 May 2020 (Day 53 after the effective lockdown), Phase Ib on 11 May (Day 60 after the effective lockdown). Phase II took effect on 18 May 2020 (Day 67 after the effective lockdown).

In this paper we analyse the relative effectiveness of the adopted measures by looking at the acceleration coefficient in the development of the pandemic. Analysing the historical data we determine the coefficients corresponding to different stages of the lockdown. Consequently, we use these coefficients and their relative behaviour to project possible scenarios of development in the coming months.

The authors would like to thank Dr Erkinjon Karimov for discussions.

### 2. COVID-19 Model

Our analysis is based on the simple model proposed in [2] for the projective analysis of the pandemic. The analysis carried out there for March 2020 yielded good projection results. We adapt it allowing a more flexible modelling of the acceleration coefficient *Rc_k_*. This coefficient takes care of the measures introduced by the government and affects the development of the infection curve. It is an accumulative quantity taking into account a variety of factors (the set of imposed rules and how strictly people follow them, quarantine measures, social distancing rules, gathering and shopping arrangements, police controls, etc.) Certainly, the considered model is simple, but it allows one to numerically measure the effects of the introduced measures and their realisation at different stages of the pandemic.

Thus, we consider

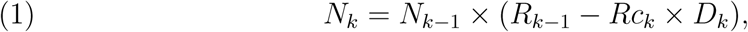

with the ratio given by

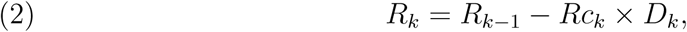

where *D*_*k*_ is the number of days between the iteration moments *k −* 1 and *k, N*_*k*_ is the total number of infected cases till time moment *k, R_k−_*_1_ is the case increase rate prior to time moment *k, Rc*_*k*_ is the function of reduction coefficient of daily cases increase rate, and

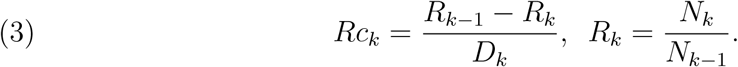

We will also carry out the similar analysis for actively infected people, without taking into account the previous history of the total number of past infections. Taking the infectious period to be 15 days, we introduce the variable

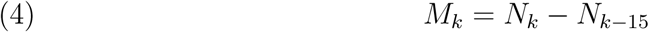

of actively infected people, without taking into account the past history of infections dating more that 2 weeks in the past. We consider for it the same equation

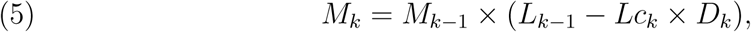

with the ratio given by

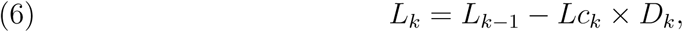

where *D*_*k*_ is the number of days between the iteration moments *k* − 1 and *k*, *M*_*k*_ is the total number of active infected cases at the time moment *k*, *L*_*k*−1_ is the case increase rate prior to time moment *k*, *L_c_k__* is the function of reduction coefficient of daily cases increase rate, and

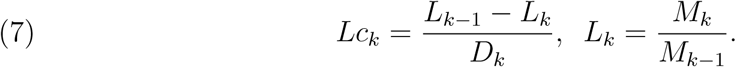

For the modelling, we put *M*_*k*_ = *N_k_* − *N*_*k*−15_ for all *k* > 15 and *M*_*k*_ = *N*_*k*_ for all *k* ≤ 15.

The second model (5)-(7) may be more suitable for modelling the development of the active phase of the infection as the number of infections of more than 2 weeks in the past has a limited effect on the development of the situation in the present.

For the simplicity and due to the daily available statistics, in both models we put *D*_*k*_ equal to one day. In what follows, we take the data in 7-day average values to eliminate inaccuracies due to weekend and other similar effects. So, the values of *N*_*k*_ are taken in seven-days averages for each *k*. Moreover, to have a clearer averaged picture we analyse the 7-day averaged values of *R_c_k__* by denoting them *Rca* in the figures below. We do the same for the coefficient *Lca*.

The higher are the coefficients *Rca* and *Lca*, the more effective are the measures. Since the proportion is taken with respect to the increasing number of cases, the values are relative to the other days in the period, and show the dynamic of the development of the infected cases in the pandemic. In general, the larger positive constants *Rca* and *Lca* would lead to faster slowing down of the pandemic.

#### 2.1. Historical Observations

In what follows we analyse the relative dynamics of different stages of the pandemic. In particular, the effects of the taken measures are visible in the graphs after about 12 days after each set of confinement measures takes place.

Although the official lockdown in Blegium started on 18 March 2020, the Belgian National Security Council decided to move into the federal phase of crisis management on 12 March. This meant that the schools, discos, cafes and restaurants were closed, and all public gatherings were cancelled. In fact, the so-called *reinforced phase 2* started already from 10 March. Thus, for the purposes of this modelling we will take 13 March as the start day of the *effective lockdown*. For brevity, when we will refer to the lockdown (e.g. in the plots) we will mean the effective lockdown (from 13 March) rather than the formal one (from 18 March).

For the convenience of understanding the graphs, we summarise the main dates for Belgium as follows:

- Effective lockdown started on 13 March 2020, which is Day 1 in Figure 2.
- Phase Ia started on 4 May (Day 53 after the lockdown)
- Phase Ib started on 11 May (Day 60 after the lockdown).
- Phase II started on 18 May (Day 67 after the lockdown).

In Figure 1, we give the 7-days averaged plots of the total number *N* of people having had the infection, and of the number *M* of actively infected people, for the period from 13 March to 31 May 2021. The relation between *N*_*k*_ and *M*_*k*_ is given by formula (4). The data is taken from [1].

In Figure 2, the graphic of the Belgian *Rca* and *Lca* after the lockdown is given. The spot coloured in red corresponds to the *Lockdown Effect*. It shows the steep increase in the *Rca* curve around Day 12 after the lockdown, signifying the positive effects of the taken lockdown measures. The same pattern is visible in *Lca*, with the last graph in Figure 2 showing the evolution of the averaged number *Lca*. This also includes the effects of the reinforced phase 2 which started 3 days before the effective lockdown.

**Figure 1.**
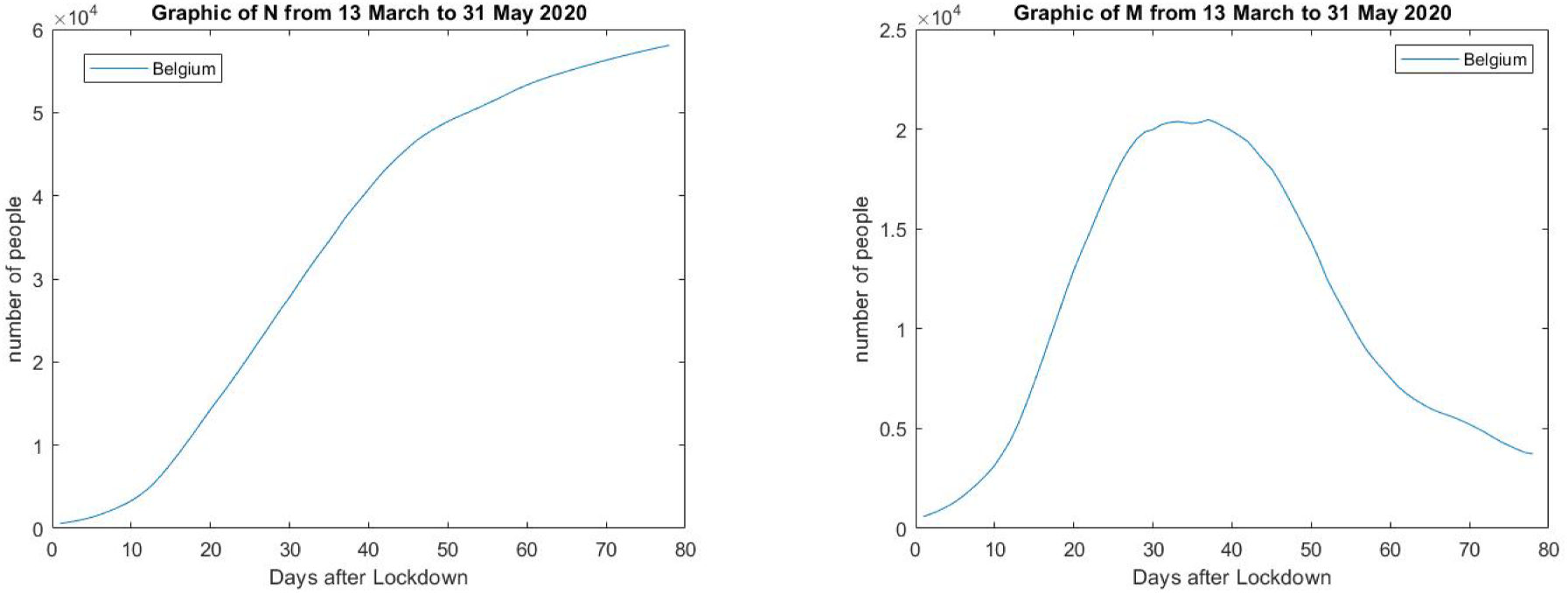
In the first plot, the graphic of the 7-day averaged number of all infected people in Belgium after the effective lockdown is given. In the second plot, the graphic of the 7-day averaged number of actively infected people in Belgium after the effective lockdown is given.

**Figure 2.**
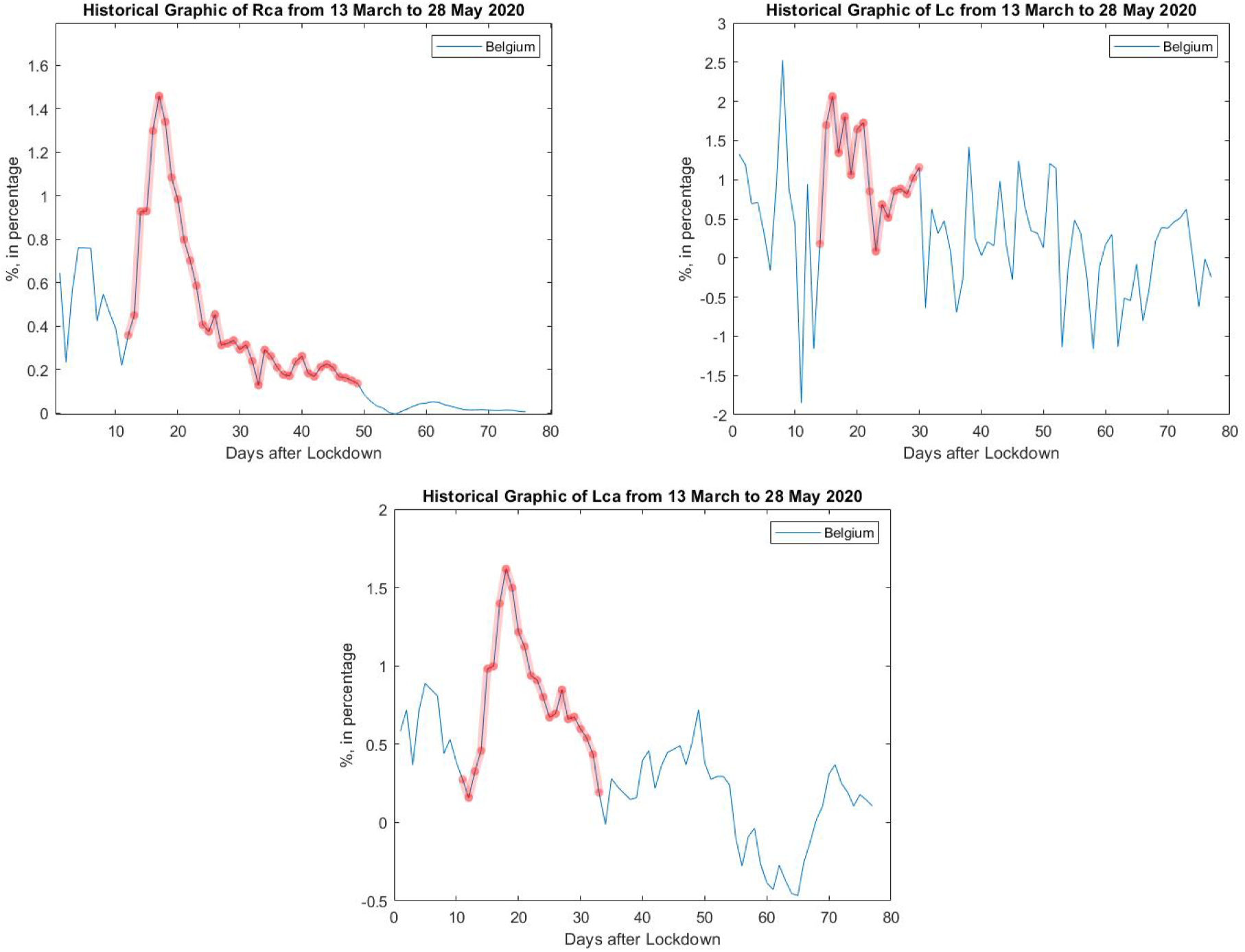
In the first plot, the graphic of Belgian *Rca* after the effective lockdown is given. The part coloured in red can be viewed as the *Lockdown Effect*. It shows the steep increase in the *Rca* curve around Day 12 after the lockdown, signifying the positive effects of the taken lockdown measures. This is also visible in the graph of *Lca*. The second and third plot show the values of *Lc* and its 7-days average *Lca*.

In Figure 3, the graphics of Belgian *Rca* and *Lca* after easing the lockdown are given. The points coloured in red show the *Phase I* and *II Effects*, respectively. They show the decreasing *Rca* signifying that the effects of the lockdown are weakened by easing the respective restrictions. The last plot shows the beginning of the evolution plot for *Lca* for Phase II based on the few days data available at this moment.

**Figure 3.**
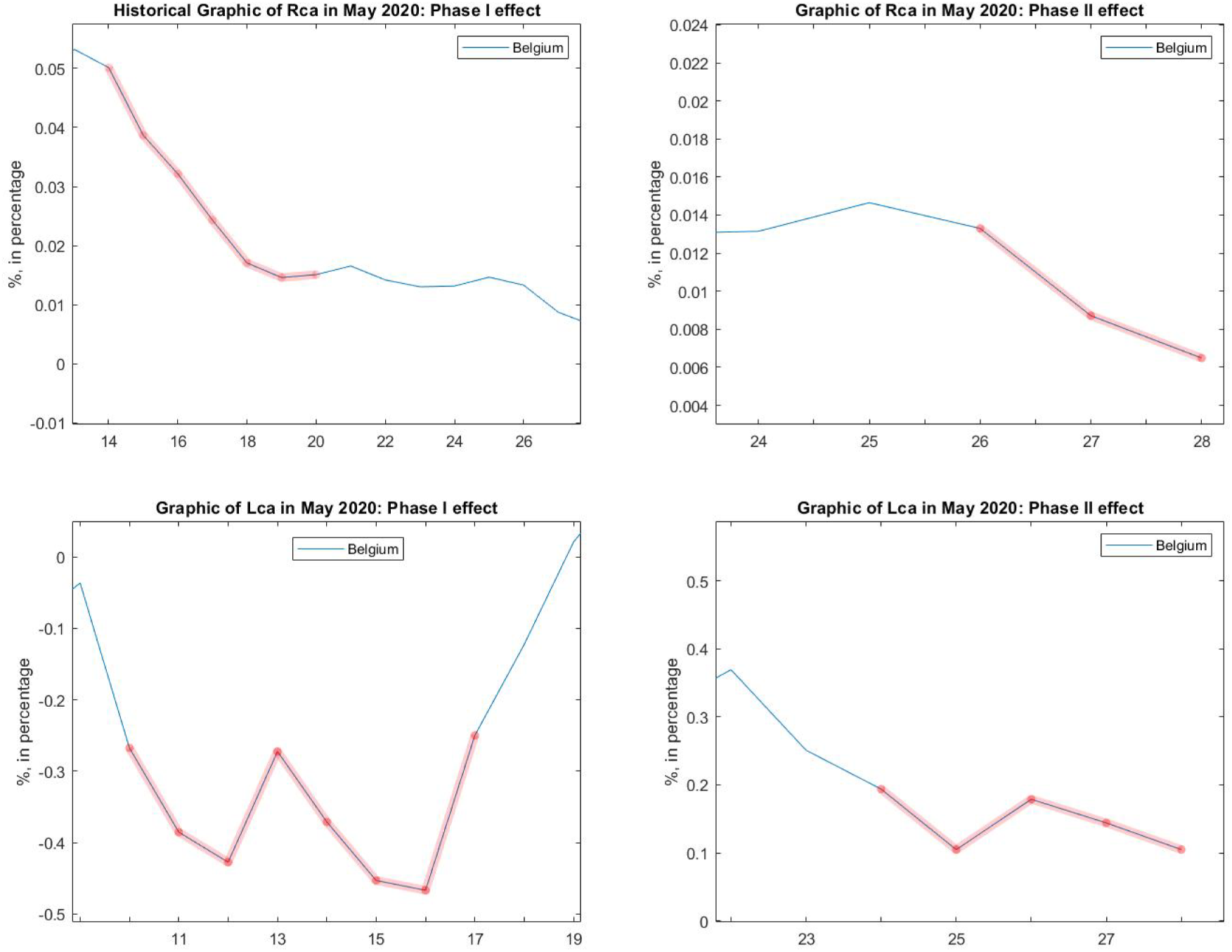
In the first two plots, the graphics of Belgian *Rca* are given after easing the lockdown. The points coloured in red correspond to *Phase I* and *II Effects*, respectively. On these figures, the numbers on the horizontal axis show the dates in May: 14-26 May and 24-28 May, respectively. The third plot shows *Lca* for the effect of Phase I which is negative relative to the previous developments; it corresponds to Days 59-66 after the effective lockdown.

In Figure 5, we see that the effects of the measure become less effective during Days 5-57 after the lockdown (the period of 1-7 May 2020). As there is a lag effect of around 12 days, this could be possibly explained by several reasons. Apart from people tiring of the lockdown measures one month after the lockdown, this can be possibly also explained by the warm weather when people are inclined going out more compared to the colder periods. The good weather shown in Figure 4 influences the period of 10-20 days, explaining the local minimum (even negative) of the value of *Rca* on 7 May 2020. The function *Lca* becomes negative in this period, signifying the relative accelerating bechavior of the number of actively infected cases.

**Figure 4.**
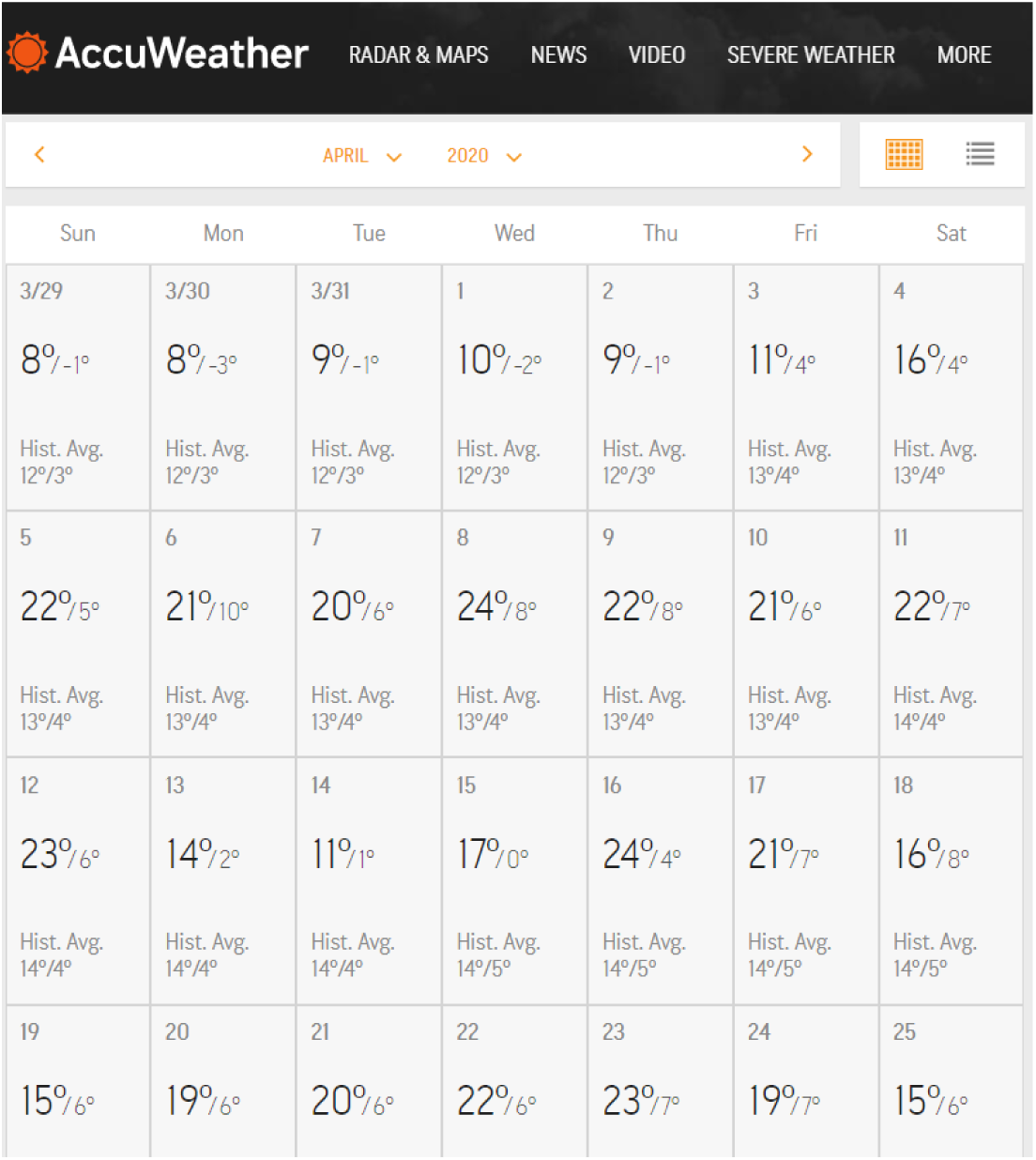
Here, information related to the weather in Brussels in April 2020 is given. The data is taken from website [3].

**Figure 5.**
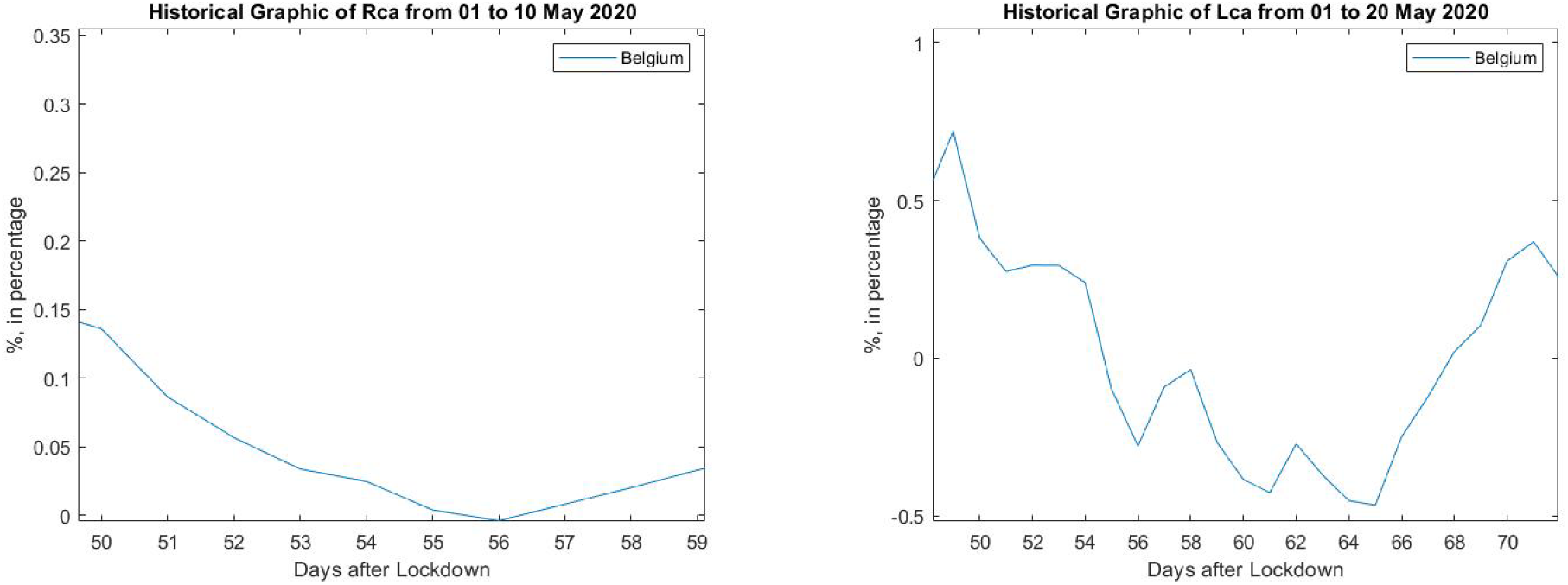
In this picture, we can see how a good weather could possibly cause problems to control the spread of the virus. The good weather shown in Figure 4 is a possible influence for the period of 10-20 days, explaining the local minimum (even negative) of the value *Rca* on 07 May 2020. Day 50 after lockdown is 1 May 2020.

In Figure 6, we can see that the values of *Rca* halved during the ten days periods between 11-20 May and between 20-29 May.

**Figure 6.**
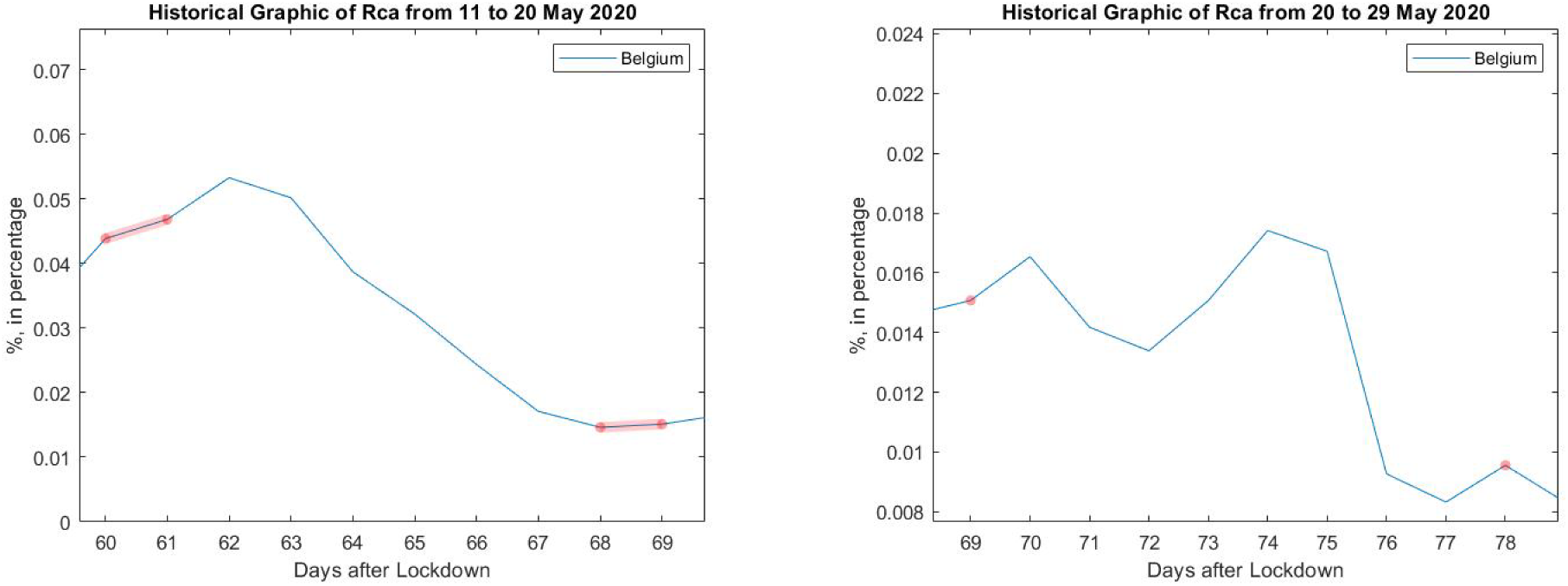
From these plots, we can see that the values of *Rca* has roughly halved in ten days period between 11-20 May and between 20-29 May. Here 11 May is Day 60 after lockdown. The picture on the left shows the decrease of *Rca* during Phase Ia, while the picture on the right shows the decrease of *Rca* during Phase Ib.

#### 2.2. Projection

In Figure 7, we give a projection for three different values of *Rca* starting from the end of May 2020. Here, we are forecasting the cases for *Rca* = 0.02, 0.015, 0.005 percentages. We observe that the scenarios with *Rca* = 0.02 and *Rca* = 0.015 are predicting the end of the pandemic in 20-30 days. But the scenario with *Rca* = 0.005 is predicting the end of the pandemic in 2-3 months.

**Figure 7.**
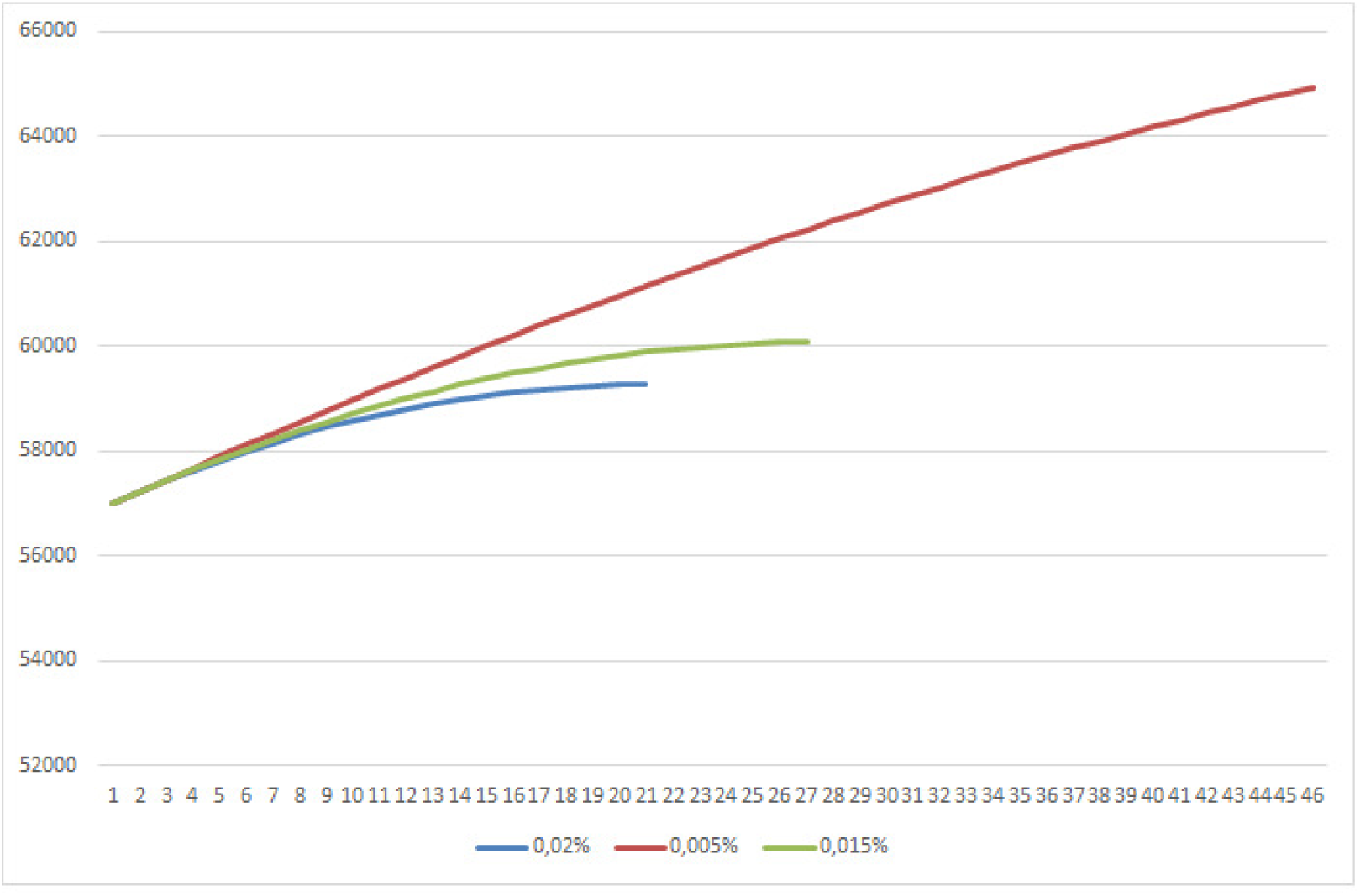
In this plot, we give a projection for three different values of *Rca* starting from the end of May 2020. Here, we are forecasting the cases for *Rca* = 0.02, 0.015, 0.005 percentages. We observe that the scenarios with *Rca* = 0.02 and *Rca* = 0.015 are predicting the end of the pandemic in 20-30 days. But the scenario with *Rca* = 0.005 is predicting the end of the pandemic in 2-3 months (this is not visible on the graph due to the scale but follows from the modelling data).

In Figure 8, we give a projection of the case with the value of *Rca* = 0.015 percentage at the end of May 2020. Here, we look at what happens when the value of *Rca* is halved over the periods of every 10, 15, and 30 days, respectively. This halving condition is natural to consider in view of the data observed in Figure 6. Thus, we are forecasting the scenarios when *Rca* is halved every 10, 15, and 30 days, respectively. We observe that the scenarios with 15 and 30 days halving of *Rca* are predicting the end of the pandemic in 20-40 days whereas the scenario with 10 days decreasing *Rca* is predicting that the pandemic could last for years.

**Figure 8.**
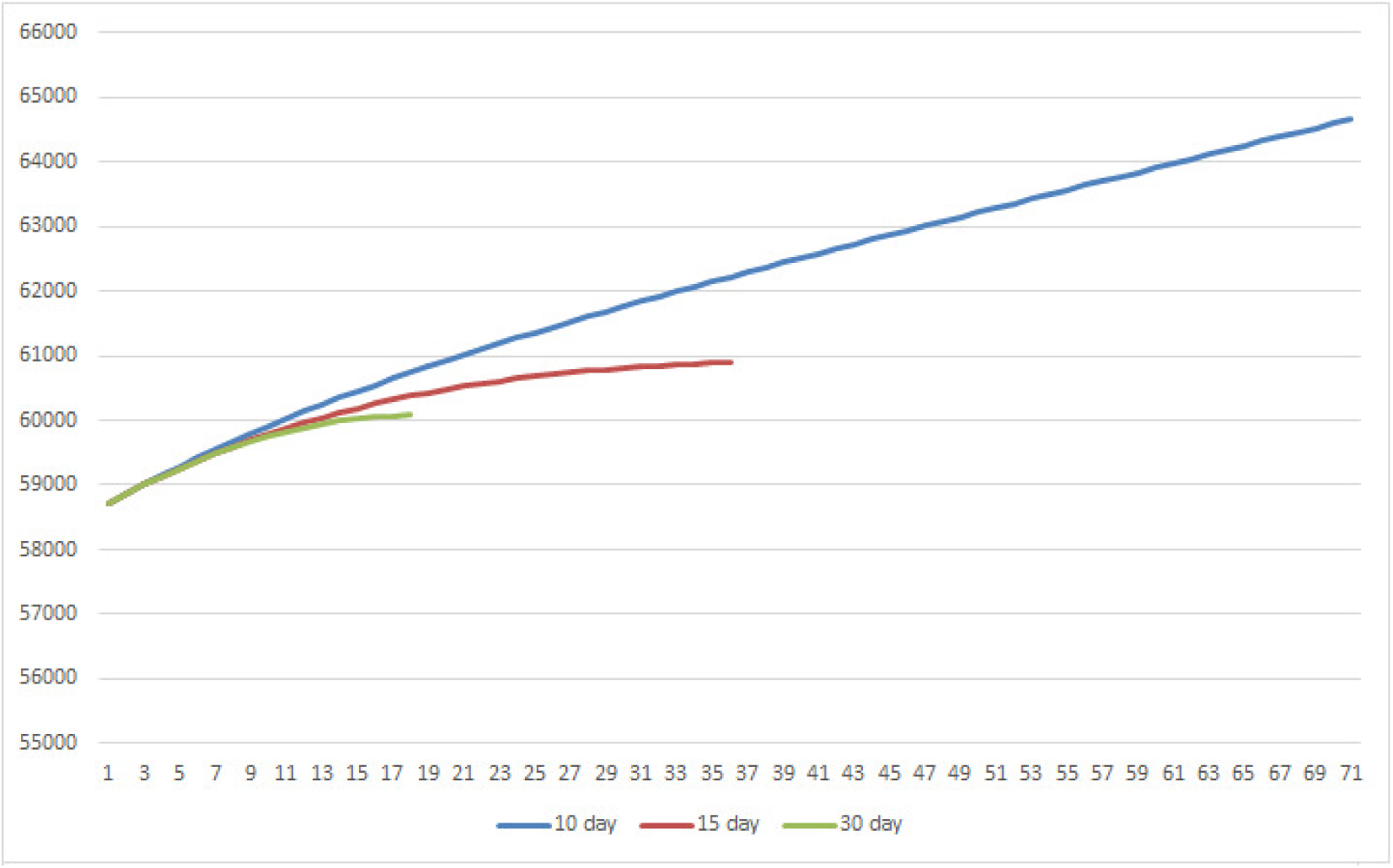
In this plot, we give a projection of the case with the value of *Rca* = 0.015 percentage at the end of May 2020. Here, we are forecasting the cases when *Rca* is halved every 10, 15, and 30 days, respectively. We observe that the scenarios with 15 and 30 days periods for halving *Rca* are predicting the end of the pandemic in 20-40 days whereas the scenario with 10 days halving *Rca* is predicting that the pandemic could last for years.

### 2.3. Recommendations

We see that as of the end of May 2020, the current *Rca* value of 0.015 keeps the pandemic manageable, in the ideal situation forecasting that the situation will be under control within a month. Further relaxation of the confinement measures is still possible, allowing one to still keep the situation under control. For example, the further reduction of *Rca* by the factor of 1*/*3 to 0.005 would still keep the situation manageable, however, extending the period of bringing the pandemic under control to up to 3 months. It is difficult to express such reduction of *Rca* in terms of the concrete measures, as it depends on a combination of factors such as the introduced government measures, their enforcement, and how closely the population follows them. However, further substantial relaxation of the confinement measures may be dangerous as it could extend the reduction period by many more months or to lead to the second wave of infections.

### 2.4. Conclusion

Using a simple model, we can analyse the relative effects of different stages of the lockdown on the numbers of infected cases. The analysis of *Rca* shows an immediate positive effect starting at 12 days after the effective lockdown (which may be 15 days if the reinforced phase 2 is also taken into account). However, a month later the measures lose their strength, possibly due to people tiring of the lockdown, and the effects of the warm weather. We see the loss of the effect of the measures after each subsequent phase, however, the effects are still strong enough to control the pandemic in principle within a month (in the ideal situation). However, it is clear that further relaxation of measures have to take place (for example for the reasons of the Economy), and that it has to be done in a controlled way. For the future projection, we fix a constant *Rca* which will be determined by further measures taken by the government. We see that a further decrease of *Rca* from 0.015 to 0.005 would extend the potential control of the pandemic (in the ideal situation) to up to 3 months. However, if the relaxation of measures continues by halving *Rca* every 10 days, the duration of the pandemic could last for years, that is, until the herd immunity is reached, or until the further spread of COVID-19 is treated by a vaccine.

## Data Availability

public data

## References

1. https://www.worldometers.info/coronavirus/

2. Jian Lu. A New, Simple Projection Model for COVID-19 Pandemic. https://www.medrxiv.org/content/10.1101/2020.03.21.20039867v2

3. https://www.accuweather.com/en/be/brussels/27581/april-weather/27581

